# Determinants of hand hygiene behaviours in internally displaced persons camps in flood-affected areas of South Sudan: a mixed-methods formative research study

**DOI:** 10.1101/2025.02.14.25322287

**Authors:** Chol Bak, India Hotopf, Zack Wol, Fiona Majorin

## Abstract

In addition to longstanding conflicts, South Sudan has experienced devastating flooding in recent years, resulting in an ongoing humanitarian crisis. In early 2024, it was estimated that there were two million internally displaced persons (IDPs) in South Sudan, with camp residents amongst those most vulnerable to infectious disease outbreaks. Our aim was to understand determinants of hand hygiene behaviours and explore existing hygiene programs in IDP camps in flood-affected areas of South Sudan. A mixed method approach was used to collect data in four IDP camps in Bentiu county, Unity State, in May to June 2023. Methods included: key informant interviews (n=5) with water, sanitation, and hygiene (WASH) implementing organizations, tours of WASH infrastructure in camps, focus group discussions (n=12) with camp residents disaggregated by age, gender, and disability status. Lastly, 50 camp residents were asked to participate in a brief survey and to demonstrate their handwashing behaviour in their current situation and in a hypothetical scenario if less water was available. We found that camp residents mostly washed hands with water only, with some using soap when available. Women were perceived as more likely to wash their hands. Residents commonly washed their hands after visiting the toilet, and before and after eating. Drivers of handwashing included visibly dirty appearance, smell, or a desire to feel fresh and clean. The main barriers to handwashing included a lack of soap, water, buckets, and other handwashing materials. Flooding affects handwashing behaviours in multiple ways, such as increased pressure on limited facilities due to IDP influxes and submerged water points. Handwashing using potentially contaminated flood water was also reported. Extreme weather events such as floods are predicted to increase in frequency and intensity with climate change - thus finding ways to sustainably improve handwashing in these changing contexts is essential.

## 1. Introduction

Hand hygiene is among the most effective behaviours that can be adopted to prevent infectious diseases, including diarrhoeal diseases and respiratory diseases (CDC, 2022; WHO, 2022a). Meta analyses demonstrate that good hand hygiene reduces the risk of diarrhoeal disease by 23-48% (Cairncross et al., 2010; Freeman et al., 2014; Wolf et al., 2022) and is a significant protective measure for cholera (Jones et al., 2020). Whilst the efficacy of hand hygiene is recognised, knowledge does not always translate to adherence. Indeed, Wolf et al. (2023) estimated that just 26.4% (23.4-29.6) of the global population wash their hands with soap after potential faecal contact. Globally, unsafe hand hygiene is estimated to cause some 750,000 preventable deaths from diarrhoea and acute respiratory infections annually (WHO, 2023; Wolf et al., 2022). Hand hygiene is shaped by multidimensional behavioural determinants; effective interventions require an understanding of the specific determinants and barriers within target settings (Aunger & Curtis, 2016; White et al., 2020a).

Humanitarian crises, including extreme weather events, are associated with infectious disease outbreaks, such as COVID-19, cholera, and diarrhoea, with diarrhoea contributing up to 40% of deaths in camp settings in acute phases of an emergency, 80% of which are among children <2 years old (Connolly et al., 2004). Crisis-affected populations typically face inadequate water, sanitation, and hygiene (WASH) access, crowded living conditions, food insecurity, and disrupted healthcare access, heightening risk (Cantor et al., 2021; Connolly et al., 2004; Lam et al., 2015; Owoaje et al., 2016).

Extreme weather events have caused 115 million deaths in the past 50 years, of which, 90% occurred in low resource settings (WMO, 2021). The number of extreme weather events have increased five-fold over this period, and the frequency and intensity is predicted to increase with climate change (WMO, 2021). By 2050, it is estimated that there will be 216 million IDPs due to climate change (UCL, 2022; WMO, 2021).

Over the past two decades, flooding has been the most common natural disaster, affecting 2.3 billion globally, and is considered the deadliest, accounting for 43.5% of natural disaster deaths in 2019 (CRED, 2020a, 2020b). Floods damage WASH infrastructure, and wastewater overflow is common and can result in contaminated water supplies (ACAPS, 2017; Confalonieri et al., 2018; Mora et al., 2022; Semenza et al., 2012). Flooding typically results in mass migration to informal settlements and camps, characterised by crowded conditions and inadequate WASH access, impeding hand hygiene (Challa et al., 2022; OCHA, 2022a). South Sudan is among the five countries most vulnerable to climate change, with the least response capacity, globally (IOM, 2024). The country has faced four consecutive years of flooding, impacting between 750,000 and one million people annually, half of whom are internally displaced (UNHCR, 2023). As of May 2023, when data for this study was collected, there were approximately 9.4 million people in need of humanitarian assistance in South Sudan, including 2.2 million IDPs due to natural disasters and conflict, with numerous ongoing cholera outbreaks (UNICEF, 2023b).

Despite the evident importance of hand hygiene interventions in populations affected by floods, the recent COVID-19 pandemic, and the fact that flooding events are predicted to increase in frequency and severity, there is a dearth of studies exploring hand hygiene behavioural determinants during, or in the aftermath, of severe flooding. Even when considering hand hygiene behaviour studies within crisis affected settings broadly, at the time of conducting this study, most evidence adopted a survey-based approach, failing to fully elucidate the complex phenomenon of behavioural determinants (White et al., 2022).

Using behaviour change frameworks ensures that behavioural determinants are considered during formative research and intervention design. This study was guided by the Behaviour Centred Design (BCD) approach, which draws on evolutionary psychology and outlines a ‘design-thinking process’, composed of five steps to design an effective behaviour change intervention: assess, build, create, deliver and evaluate (Figure 1) (Aunger & Curtis, 2016). This formative study pertains to the ‘assess’ and ‘build’ steps of the process, which will ultimately feed into the design and implementation of a tailored behaviour change intervention.

**Figure 1.**
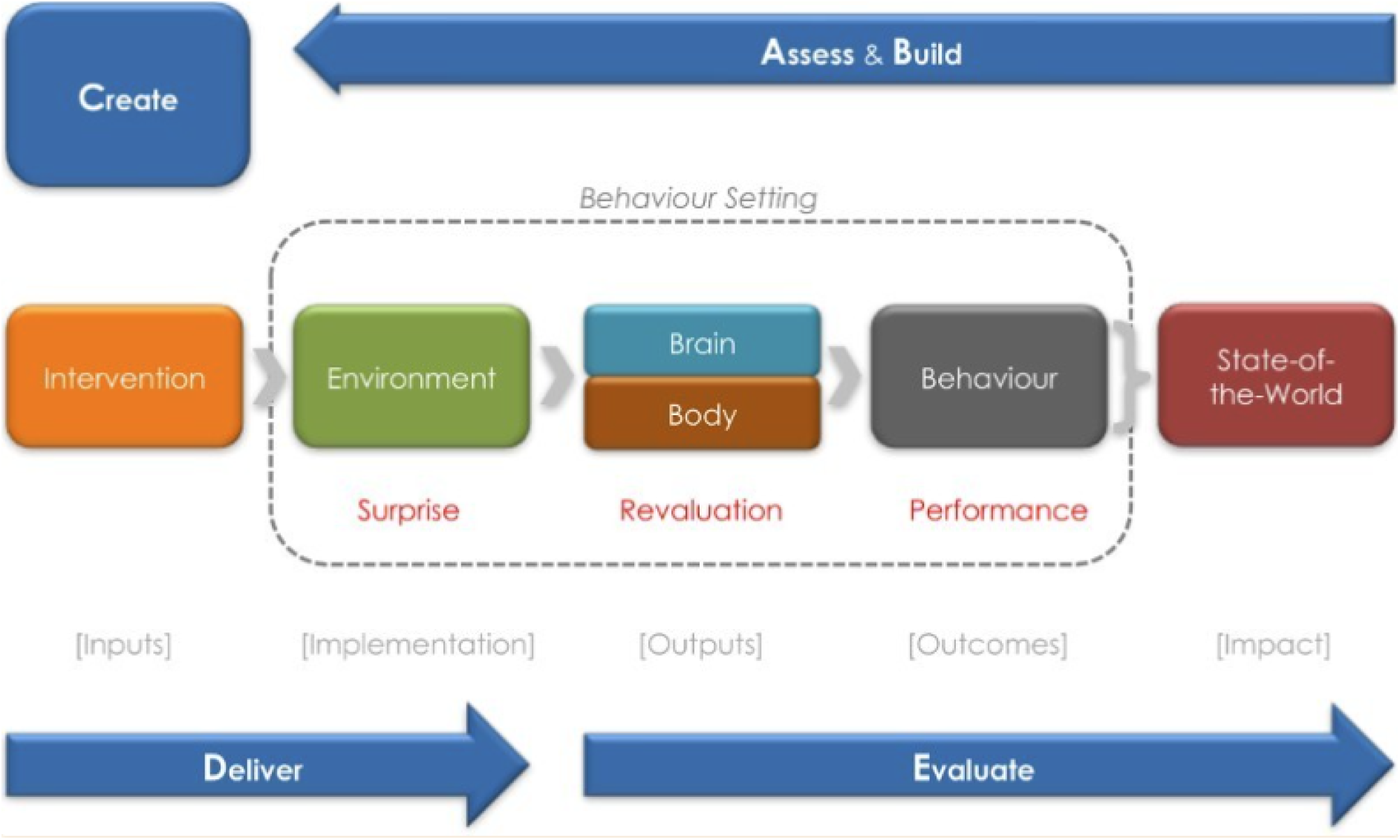
BCD Framework. The sections in blue represent the five key steps, whilst the theory of change is portrayed across the middle. The first step involves ascertaining what is known and unknown about the focal behaviour and its determinants and the second step entails addressing identified knowledge gaps through conducting formative research. Next, researchers design a behaviour change intervention, from the concept to specific materials and activities to be implemented. Once designed, the intervention must be delivered and evaluated to determine whether the “environmental, psychological and behaviour changes” have taken place (Aunger & Curtis, 2016)

**Figure 2.**
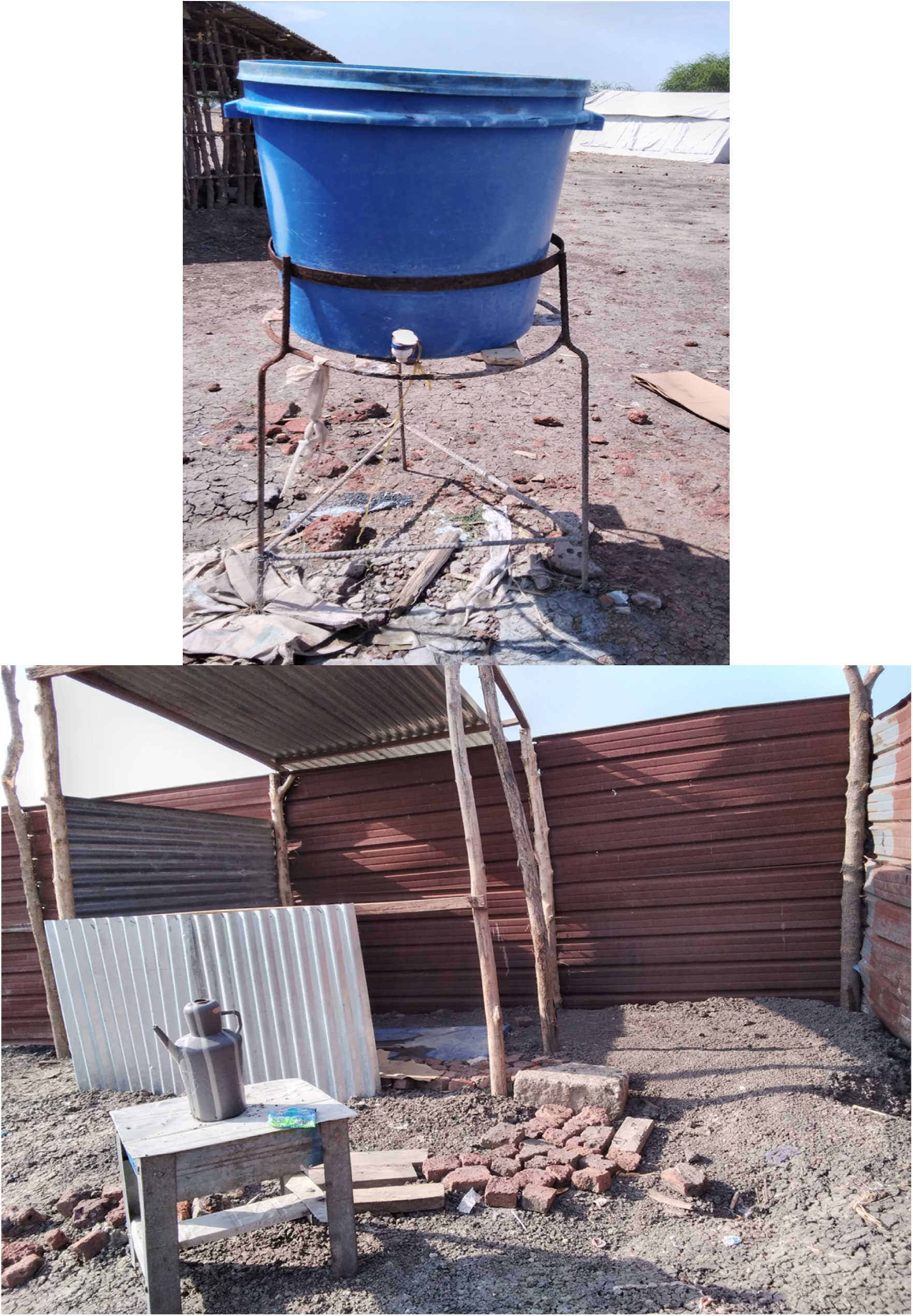
Images of some handwashing facilities taken during camp tours. Top shows a bucket with a tap, bottom shows an ibrik on a table outside a latrine (the second would not have counted as a permanent handwashing facility in the demonstrations).

**Figure 3.**
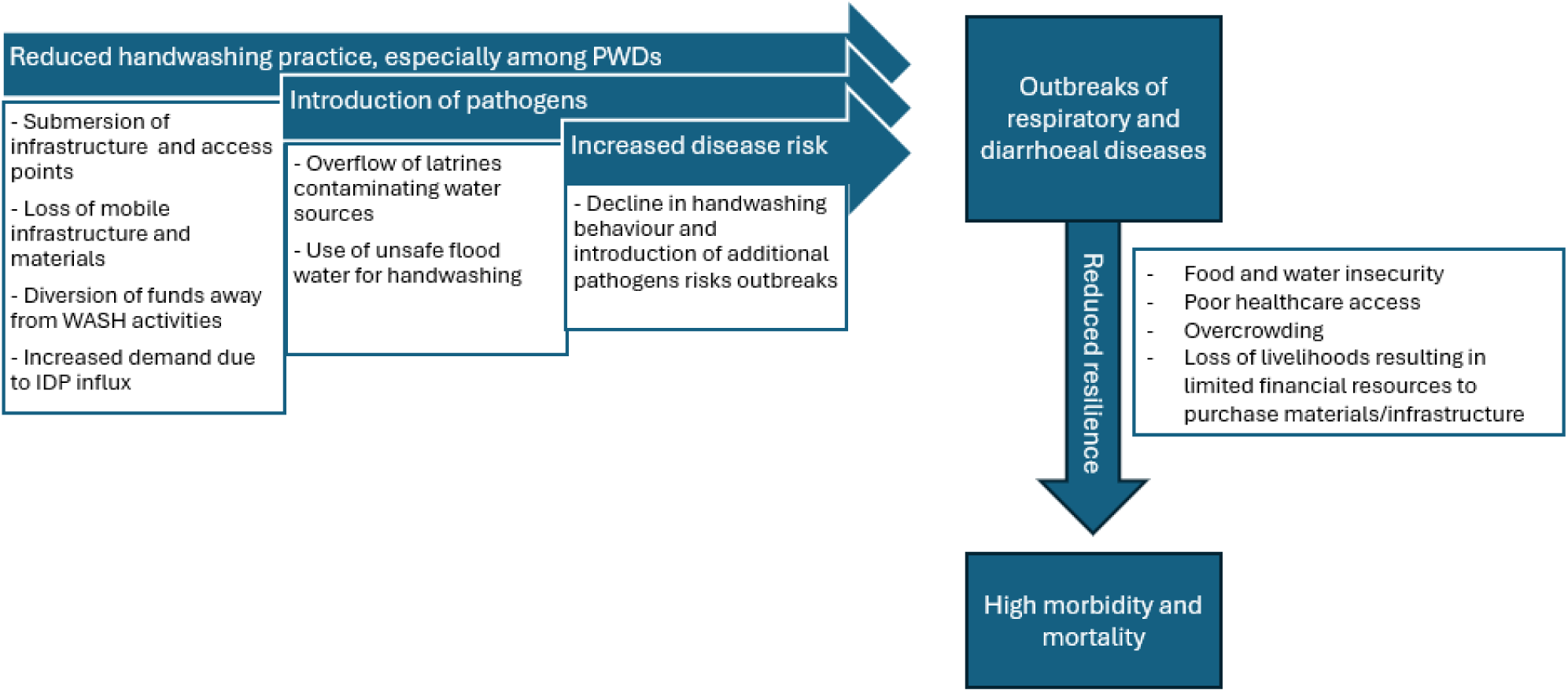
Multifaceted impacts of flooding (original figure)

This study aimed to understand the determinants and barriers to hand hygiene behaviours and explore existing hygiene promotion programmes in internally displaced person (IDP) camps in flood-affected areas of South Sudan, through 1) exploring existing WASH programmes and challenges in delivery and 2) elucidating behaviour determinants and barriers to hand hygiene within the context of flood-affected IDP camps.

## 2. Materials and Methods

This formative research study used mixed-methods and engaged crisis-affected persons, IDP camp managers and WASH professionals through hand washing (HW) demonstrations, key informant interviews (KIIs) and focus group discussions (FGDs) using participatory activities (see supplementary material A for an overview of the methods in relation to BCD determinants). The study was conducted in parallel with a study in Somalia, looking at determinants of hand hygiene in IDP camps in drought-affected areas.

### 2.1 Study sites

The study was conducted in Unity State in Northern South Sudan, in 4 IDP camps in Bentiu town: the Protection of Civilians camp (POC), which is the largest IDP camp in the country, hosting 102,436 IDPs as of March 2023 (IOM, 2023), and three smaller nearby IDP camps that host flood displaced persons. Bentiu town was selected as the study setting, as it has been severely affected by floods and has camps hosting flood displaced populations.

### 2.2 Participant sampling and data collection

The research team delivered training on research methods and co-designed standard operating procedures (SOPs). Four research assistants (RAs) were recruited from the Bentiu community (two male, two female). On May 9^th^, RAs received in-person data collection training, and data collection occurred from May 15^th^ –25^th^ June 2023. Camp tours were conducted first (n=4), followed by the HW demonstrations (n=50) and FGDs (n=12). KIIs were conducted in parallel (n=6).

#### Camp tours

Camp tours were conducted in each camp to understand available WASH infrastructure and challenges. Camp managers were approached and asked to provide guided camp tours after verbally consenting to participate in the study. Hand drawn maps created on these tours were then used for reference during the FGDs. Finally, short interviews were conducted with camp managers in Nuer, with field notes and audio recordings taken.

#### Handwashing demonstrations

50 participants were randomly selected to conduct a handwashing demonstration using an adaptation of the Extended Program of Immunization (EPI) sampling method (Bostoen & Chalabi, 2006). 25 were recruited from the largest camp, with even distribution across remaining 3 camps. Prospective participants received study information and provided written informed consent. There was an even split of women and men.

Demonstrations were based on the Wash’Em tool and entailed asking participants to demonstrate how they usually wash hands before preparing food (women) or after the toilet (men and women) (Wash’Em, 2023). Next, they were asked to demonstrate how they would wash their hands if water was scarcer. Soap was not mentioned to avoid bias. Each participant completed two demonstrations, totalling 100 demonstrations.

Demonstrations were video recorded using mobiles and observations on participants’ interactions with infrastructure, materials (including presence of soap or soap alternative), water and suitability of the environment, were captured on tablet Kobo (KoboCollect, Cambridge, Massachusetts, United States) forms. Additionally, comments were noted. Finally, a short questionnaire on handwashing practices and materials was conducted using a Kobo form.

#### Focus group discussions

12 FGDs were conducted with eight participants each. Participants were recruited from across the four camps, with maximum variation for age, sex, and disability status. To minimise potential bias introduced by hierarchal structures, FGDs were disaggregated according to age, gender and disability status (**Error! Reference source not found.**). Note that only individuals with physical disabilities were included.

Recruitment was facilitated by camp managers, who engaged residents via existing community groups e.g. disability groups. Camp managers were provided with study information, including information sheets and the Washington Group questions to facilitate recruitment of persons with disabilities (WG, 2022). Camp managers used the exclusion criteria and sampling frame to identify eligible participants, before reading the contents of information sheets and asking for permission to share participants’ contact details with researchers to arrange FGDs. Children, non-IDP residents, people with intellectual disabilities and people who did not consent were excluded.

On the day of FGDs, verbal information on the study and FGD procedure was provided again in Nuer. After checking understanding, participants provided verbal consent. RAs worked in groups of two (one male and one female), alternating between facilitator and note taker.

FGDs were guided by topic guides. Two topic guides were created in relation to BCD determinants: one explored behaviour, infrastructure, and materials and the second related to norms, motives and knowledge. Both included questions about the impacts of flooding. FGDs were conducted in Nuer and audio was recorded. Topic guides were iteratively refined as new findings emerged (supplementary material B).

We planned to conduct 16 FGDs (two per participant type) but stopped after 12 as the research team felt that saturation had been reached, with no new information emerging (Table 1).

**Table 1.** Sampling frame for FGDs.

| **Participant type** | **Definition** | **n** |
| --- | --- | --- |
| Young women | 18-25 years old | 1 |
| Young men | 18-25 years old | 2 |
| Older women | 26 – 64 years old | 2 |
| Older men | 26 – 64 years old | 1 |
| Elderly women | 65 + years old | 2 |
| Elderly men | 65+ years old | 2 |
| Women with disabilities | Self-identified physical disability (all ages) | 1 |
| Men with disabilities | Self-identified physical disability (all ages) | 1 |

#### Key informant interviews

Six KIIs were conducted with five males and one female, with a mean length of 37.41 minutes (range: 9:50 to 59:58 minutes). To recruit participants, researchers engaged the national WASH Cluster lead who made introductions with NGO organisations operating in the camps in Unity state. In turn, organisations suggested potential respondents who were invited for interviews. Prospective participants received information sheets in person and provided written informed consent.

KIIs were conducted in person in English, with audio recordings taken. KIIs followed a topic guide (supplementary material C) which included questions on current programmes and infrastructure; handwashing behaviour of camp residents; disease prevention and perceptions; avenues for improving handwashing and closing questions. One KII recording was not used due to recording quality issues, thus five were analysed in the end.

### 2.3 Data management

Audio recordings were translated (where required) and transcribed verbatim by BNO researchers fluent in Nuer and English. All identifiable information was removed, and participants were assigned unique identifying numbers. Anonymisation occurred at the individual and organisational level. During data collection, audio recordings were stored on computers belonging to partner organisations only, and files were password protected. Data was uploaded to Microsoft SharePoint (a secure platform), access was restricted to researchers and previous copies were deleted. During data collection, there were debriefs conducted between LSHTM and BNO researchers to discuss reflections, challenges and topic guide amendments.

### 2.4 Data analysis

Prior to data analysis, LSHTM delivered training on mixed-methods analysis and on using Dedoose, a cloud-based qualitative data analysis software (Dedoose, Los Angeles, California, United States). Training was also conducted on producing a deductive framework, which covered the different BCD dimensions (Aunger & Curtis, 2016).

Qualitative data was thematically analysed using the six-step process outlined by Braun and Clarke (2006). Dedoose was used to code data deductively using a unified coding framework. As inductive themes emerged, changes and comments were captured on a live coding framework and discussed in team meetings to ensure completion of coding of each transcript, intercoder reliability and rightful application of codes. CB conducted all coding and IH double coded two FGDs and KIIs in full and reviewed the rest of the coding throughout, providing guidance via regular calls to discuss challenges and iterative changes to the coding framework. CB kept a coding diary, to record thoughts and encourage reflexive practice. Data was charted in Word and summaries were produced with illustrative quotes. Once analysis was complete, the research team held a participatory analysis workshop and results were written up and synthesised under relevant BCD factors and sub-factors (Aunger & Curtis, 2016). Additionally, the Wash’Em motives exercise Excel sheet was populated (Wash’Em, 2023). Demonstration videos were watched and data was recorded in an adapted Wash’Em Excel sheet (Wash’Em, 2023) by CB. Half of the videos were also watched and recorded by FM to ensure agreement in data entry. CB conducted descriptive analysis of the survey and demonstration data in Excel.

### 2.5 Ethics and consent

Ethical approval was obtained from the LSHTM ethics committee (Ref: 28372) and the South Sudan Ministry of Health (MOH/RERB/A/05/2023). Participation in the study was voluntary, and participants were only enrolled after receiving complete details of the study in their local language and providing oral or written consent (see above). Participants could withdraw at any time without penalty and confidentiality was ensured.

There were some deviations in the consenting procedures that occurred during the data collection, which were reported to the ethics committees. These were that we had originally intended for 1) the FGD consent process to be written rather than verbal; 2) for the camp manager interviews to not be recorded and 3) for camp manager consent to be written.

The research assistants were sensitive to potential struggles that participants may be experiencing and thus made distress planning strategies ahead if the data collection such as mapping out support services, offering interview breaks or termination and monitoring signs of distress.

## 3. Results

Overall, we conducted four camp tours, six KIIs, 50 handwashing demonstrations and 12 FGDs with a total of 152 participants. Five out of the six key informants were men, whilst FGD and handwashing demonstrations were evenly split by gender (.Table 2). The mean age of handwashing demonstration participants was 34 (range: 18-57) and the mean age of FGD participants was 44.6 years (range: 18-78). FGD participants had lived in the camp for a mean of 4.9 years (range:1-9), the main religion of participants was Christianity (58.3%), followed by those who identified as not affiliating to any religion (41.7%). On average, participants had 10.6 household members, including 4.3 children under age five. Most participants had not attended formal education or had dropped out at primary level (see supplementary material D).

**Table 2.** Overview of the sample size and gender of participants for each methodology.

| Methodology | n | Females | Males | Total participants |
| --- | --- | --- | --- | --- |
| Camp tours | 4 | N/A | N/A | N/A |
| KII | 6* | 1 | 5 | 6 |
| HW demos | 50** | 25 | 25 | 50 |
| FGD | 12 | 48 | 48 | 96 |
| Total | 71 | 74 | 78 | 152 |

\*only 5 KIIs were analysed due to recording quality issues. \*\*In each HW demonstration, participants conducted two demonstrations - one in their normal circumstance and one if they had less water

### 3.1 Behaviour

During handwashing demonstrations, 32 (of 50) participants demonstrated handwashing using water only, while 16 (of 50) used soap. In the handwashing demonstration survey, all 50 participants said there were times where they only used water to wash hands. Survey participants reported they wash hands with soap between 0-10 times per day, with the majority (32/50) washing hands with soap zero times per day (see table 3). More than half of the FGDs, including all groups except for female PWDs, reported a lack of handwashing soap in the camps. Few FGDs and one key informant reported ash use as rare, with one older male also using crow soap (a local plant). Only two participants used ash during the HW demonstrations. Of the 36% (18/50) of demonstration participants to either use soap or ash during the demonstrations, 11 were women and 7 were men.

**Table 3.**
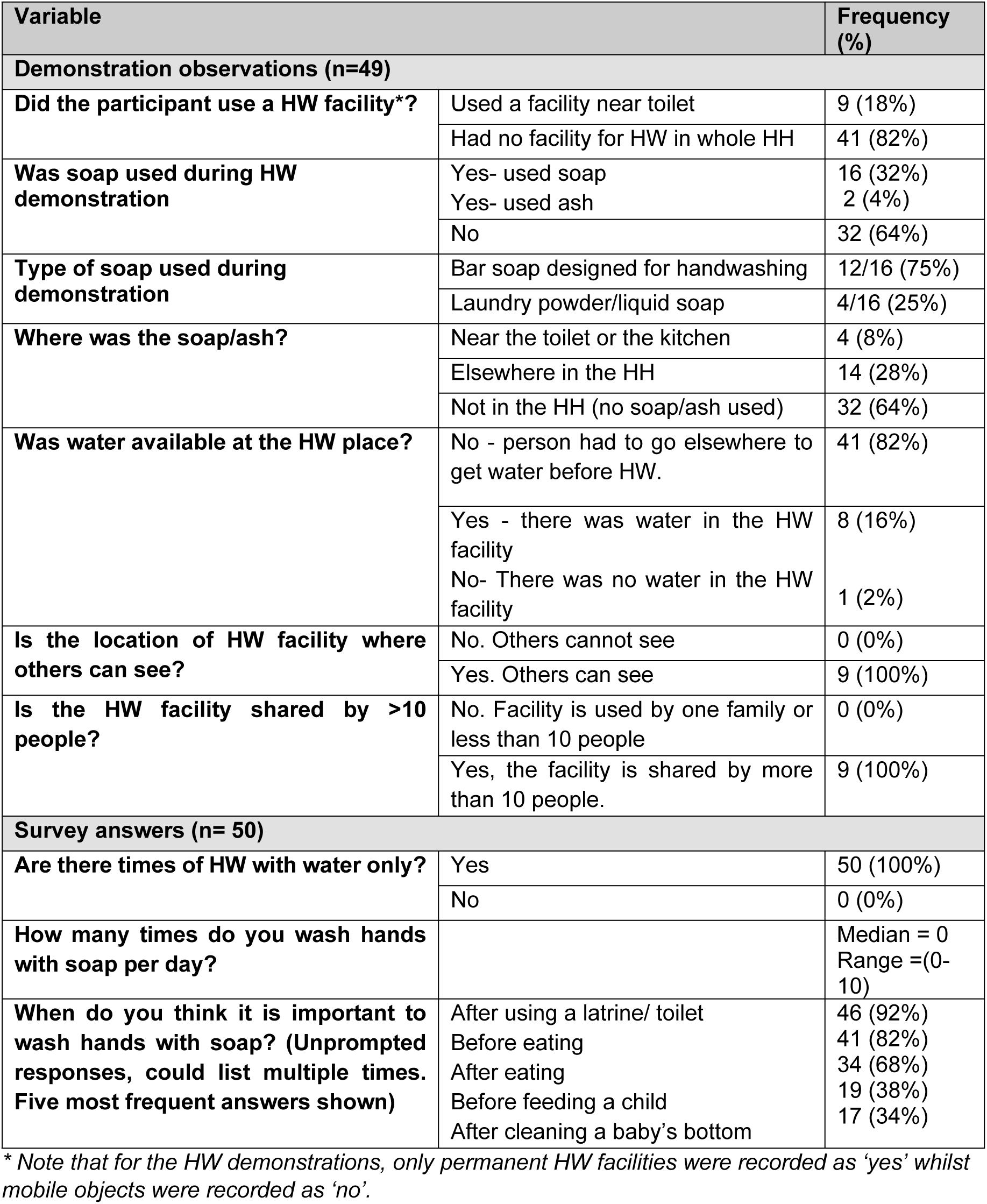
Summary of handwashing demonstration results.

| Variable |  | Frequency (%) |
| --- | --- | --- |
| **Demonstration observations (n=49)** |  |  |
| **Did the participant use a HW facility\*?** | Used a facility near toilet | 9 (18%) |
|  | Had no facility for HW in whole HH | 41 (82%) |
| **Was soap used during HW demonstration** | Yes- used soap | 16 (32%) |
|  | Yes- used ash | 2 (4%) |
|  | No | 32 (64%) |
| **Type of soap used during demonstration** | Bar soap designed for handwashing | 12/16 (75%) |
|  | Laundry powder/liquid soap | 4/16 (25%) |
| **Where was the soap/ash?** | Near the toilet or the kitchen | 4 (8%) |
|  | Elsewhere in the HH | 14 (28%) |
|  | Not in the HH (no soap/ash used) | 32 (64%) |
| **Was water available at the HW place?** | No - person had to go elsewhere to get water before HW. | 41 (82%) |
|  | Yes - there was water in the HW facility | 8 (16%) |
|  | No- There was no water in the HW facility | 1 (2%) |
| **Is the location of HW facility where others can see?** | No. Others cannot see | 0 (0%) |
|  | Yes. Others can see | 9 (100%) |
| **Is the HW facility shared by >10 people?** | No. Facility is used by one family or less than 10 people | 0 (0%) |
|  | Yes, the facility is shared by more than 10 people. | 9 (100%) |
| **Survey answers (n= 50)** |  |  |
| **Are there times of HW with water only?** | Yes | 50 (100%) |
|  | No | 0 (0%) |
| **How many times do you wash hands with soap per day?** |  | Median = 0<br>Range =(0-10) |
| **When do you think it is important to wash hands with soap? (Unprompted responses, could list multiple times. Five most frequent answers shown)** | After using a latrine/ toilet | 46 (92%) |
|  | Before eating | 41 (82%) |
|  | After eating | 34 (68%) |
|  | Before feeding a child | 19 (38%) |
|  | After cleaning a baby's bottom | 17 (34%) |

\* Note that for the HW demonstrations, only permanent HW facilities were recorded as 'yes' whilst mobile objects were recorded as 'no'.

Typically, soap is prioritised for washing clothes and dishes, or handwashing if visibly dirty, oily, or odorous e.g., after eating fish. When adequate water and soap is available, participants from demonstrations reported they wet both hands, rub soap and pour enough water to rinse their hands clean. When less water is available, residents typically resort to washing one hand only or not using soap to reduce water requirements. Of the 16 HW demonstration participants that used soap in the first demonstration, only eight used soap in the second demonstration when demonstrating handwashing with less water. They explained this was due to lack of enough water to rinse hands after using soap. During rainy season, participants reported washing their hands into buckets and pouring the water outside after the rain stops.

The survey found that most camp residents think it is important to wash hands with soap after visiting the toilet (92%, 46/50), before (82%, 41/50) and after eating food (68%, 34/50). This was followed by prior to feeding a child (38%, 19/50) and after cleaning a baby’s bottom (34%, 17/50). These same three critical time points were consistently highlighted across all 12 FGDs irrespective of age, gender, and disability status. Women and older women also reported HW before food preparation and after cleaning their baby’s bottom; this was echoed by a few young men (please see supplementary material E for the full handwashing demonstration results).

### 3.2 Brain

#### Executive brain

Camp residents are largely aware of the link between handwashing and disease prevention. More than 80% (40/50) of survey participants mentioned handwashing helps to get rid of germs, makes one clean and prevents diseases (See Supplementary material E). The main diseases of concern according to FGD participants were diarrhoea, typhoid, malaria, cholera, hepatitis, and skin diseases such as skin rashes and chicken pox. In all six FGDs where knowledge on disease prevention was explored, all groups related handwashing to prevention of diarrhoeal diseases. A couple of KII participants noted that some residents attribute disease to witchcraft or jealousy from neighbours. KII participants also felt that most camp residents understand the causes and symptoms of diarrhoeal and respiratory diseases and recognise hand hygiene as a key preventative behaviour.

> *“You may greet someone who has recently come out of a latrine, you will both shake hands without hesitation he must greet you with his bare hands while sweating. One…who has the clue about health issues must wash her/his hands before other things. That’s how we can prevent ourselves from diarrhoea, we must support people to wash their hands and then have their meals” (young man, FGD)*

Most camp residents demonstrated that they are knowledgeable about the importance handwashing at key times and are willing to wash their hands, with handwashing forming a part of their daily routine – evidenced by FGDs and handwashing demonstrations. Key informants also felt most residents are aware of key handwashing times.

Due to limited resources in the camps, female FGD participants revealed that they plan their water and soap allocations, and soap usage is mostly prioritised for purposes other than hand washing. Most male FGD participants across various age groups described washing hands when visibly dirty and prioritising soap for certain times e.g., after eating odorous meals. Young men also highlighted handwashing as desirable during courtship, since women might avoid engaging with men that do not wash their hands and perceive them as “dirty”.

When asked about their main worries since arriving in the camp, FGD participants highlighted their concerns about lack of food, water shortages, disease outbreaks, lack of handwashing materials, loss of property and lives due to more flooding, and lack of clothes. In addition, participants across 10 FGDs of different genders, age, and disability status consistently highlighted their inability to always wash hands due to limited access to water and handwashing materials, such as soap. Additional concerns included food insecurity due to the loss of cattle and income generation opportunities, exposure to disease due to crowded conditions, open defecation, and insufficient access to mosquito nets and medical care. Some were also concerned about snakebites, especially among children playing in flood water, as well as poor access to safe water (with long queues being common), clothing and the need to travel long distances if reallocated.

When asked about their sources of hand hygiene information since joining the camp, 7 survey participants reported they didn’t receive any information about HWWS since arriving in the camp. Of the 43 who did, 81% reported receiving information from hygiene promotors, 79% from organisations working in the camps, 58% from radios, 53% from health care workers and 5% from posters. The sources deemed the most trustworthy by participants were organizations working in camps (51.2%, 22/43), hygiene promoters (44.2%, 19/43) and health workers (4.7%, 2/43).

#### Motivated brain

In the two FGDs that completed the motives exercise, wherein participants ranked 13 characters (e.g., “a person who is a good parent” or a “a person who is wealthy and has a nice house”) based on how likely they were to always wash their hands with soap, the key motives for handwashing were the desire to be neat, the desire to be comfortable, the desire to be a good parent (nurture) and status/ wisdom.

#### Reactive brain

According to the FGD participants, most people habitually wash their hands and face early in the morning. Some camp residents said they do not wash hands before eating if they use spoons. Handwashing before eating is practiced when eating with one’s bare hands, which is typically the norm. Participants also said they wash their hands when they appear visibly dirty.

### 3.3 Body

#### Characteristics

People with physical disabilities face unique HW barriers – for instance, they are unable to reach some water points, including the main camp water tank, due to muddy paths. Consequently, some PWDs, especially older individuals, reported begging for water. This hindered their ability to wash their hands frequently.

> *“Well, we the people living with disabilities are not able to get water most of the time, I don’t reach [the] main tank, I just beg those who are in the first line to give me some water” (PWD woman, FGD)*

#### Senses

FGD participants highlighted that handwashing makes one feel fresh and nice and makes people more receptive to greet them with a handshake, the traditional mode of greeting. People prioritise handwashing with soap if meals are odorous (e.g., fish) or leave their skin oily or visibly dirty.

### Capabilities

FGD participants found it difficult to wash hands during mid-day as it is very hot and there is not enough water available. Handwashing was also difficult during big ceremonies, such as weddings, as there are many people and not enough water for all to wash hands.

Several FGD participants mentioned that new IDPs generally lacked knowledge on hand hygiene and toilet use and were perceived as less likely to wash their hands.

> *“Okay, mid-day is the worst time because it is very hot, people stay indoors, water to drink will not be enough due to hotness, people do go under shades of trees, you cannot walk whenever you wake up at your very first steps” (Older man, FGD)*

### 3.4 Behaviour setting

#### Stage and infrastructure

Handwashing takes place within households and public handwashing facilities, with the former being most frequent. Of the 50 handwashing demonstration participants, most (82%) did not use a dedicated handwashing facility. The 9 (5 males and 4 females) participants using a dedicated handwashing facility, demonstrated after using the latrine and used facilities near the latrine (mostly bucket with taps, previously installed by NGOs), 8 of the 9 facilities had water in them at the time of demonstrations, but only 4/9 had soap at the site. There were no demonstration households with permanent handwashing facilities at home; handwashing is done within any area of the compound using mobile objects, mostly *Ibrik* (plastic kettle), jugs, jerrycans and drinking water cups. In households, all participants had to obtain water from elsewhere to wash hands at the handwashing place, since most handwashing materials were mobile objects that do not store/keep HW water. FGD participants also reported they mostly use *Ibrik*, jerrycans or a bucket with a tap for handwashing.

Five FGDs of different gender, age and disability status deemed handwashing infrastructure to be insufficient – echoed by a few key informants. In some FGDs and KIIs, participants mentioned that some public handwashing facilities have been vandalised by children or stolen by camp residents. Two KII participants highlighted inadequate facility maintenance following installation by the NGOs, owed to poor management of facilities by residents and management.

In the demonstrations, all nine participants who used dedicated handwashing facilities, were only able to wash one hand a time due to the inconvenient design (button to press for water). In one FGD, participants reported that the design of public water points made it difficult to wash hands directly from the tap. Moreover, the handwashing facilities observed in the demonstrations were not attractive (e.g., aesthetically pleasing, with mirrors or decorations, and clean). All (n=9) handwashing facilities used in the demonstrations were shared by more than 10 people and were located where others could easily see them.

> *“we don’t have soaps for washing our hands and we don’t have plastic kettle to carry water for washing, so we wash near shelter […] but we don’t have plastic kettle to carry water even small water drinking cup is not there, and if you use that small black cup it can fall inside the water container, those small cups are not enough, no one is providing them, the cups for drinking water which we use to fetch water for drinking are not available, they are really not enough” (older woman, FGD)*

#### Props

68% (34/50) of demonstration participants had no soap for handwashing at the handwashing place, while 28% (14/50) had soap/ ash kept elsewhere in the house and 8% (4/50) used soap at the handwashing facility near the toilet. FGD participants also mentioned the use of soap for handwashing was uncommon. They said that hygiene kit distribution has waned, with one participant asserting a bar of soap costs 2000 SSP (South Sudan Pounds) (∼4 USD in May 2023 USD). Thus, many resorted to washing their hands with water only, or prioritising soap for other activities. Where participants did have soap, it was typically bar soap, this was also seen in the demonstrations (12/16).Some also used soap alternatives (see 3.1 Behaviour).

> *“We use ash, water and soap for washing our hands, but it depends on the situation. If you cannot afford soap, you use ash. Ash is a common material for hand washing since it is found locally in camps. Toilets are washed with water if there is no soap. Hands gloves and detergents and sanitizer is not available too.” (older man, FGD).*

#### Roles, routine and norms

In FGDs it was consistently mentioned that mothers, responsible for childcare, food preparation and water collection, were expected to wash hands frequently. One key informant mentioned school children were likely to wash their hands more often than adults since they are more receptive to handwashing messages.

For most camp residents, normal handwashing behaviours include washing hands and face first thing in the morning after waking up. For both men and women, FGD participants’ daily routines include washing their face, legs and brushing their teeth in the morning. They also eat food throughout the day, rest, visit latrines, wash their hands before eating and after visiting the latrine. Both women and men described engaging in livelihood activities, such as farming, fishing and attending to small businesses e.g., shops.

There were also gendered differences in routines; men erect the family shelter during the day, cultivate the garden and play dominos in the market area with friends, as well as resting more than women. Women’s routines include family food preparation, collecting household water, water lily collection and cleaning, weeding the garden, childcare, caring for the sick and cleaning the household – thus, the community perceives them as washing their hands more regularly. It is stigmatised for men to help women do household tasks like cooking. Unlike women, traditional and religious leaders are not expected to observe strict handwashing behaviours by virtue of their roles. Handwashing and cleanliness are also considered important among young men, as perceptions of being unclean may repulse women. Handshaking is the norm of greeting in the camps, but people are reluctant or unwilling to shake hands with individuals with visibly dirty hands or those perceived to be dirty people.

> *“How can we abandon handwashing, we wash because we fear being buried dirty when we died, if you have bad body smell, your family will refuse to prepare your food, bathing is God made, God told people to wash their hands.” (male PWD, FGD)*

Young participants mentioned they go to school as part of their daily routine, whilst PWDs described staying home and helping baby sit their children whilst their partner searches for household food.

> *“I am disabled who have nothing to do much, since I woke up yesterday eh, I was sitting home, as a babysitter, because the mother of the baby went and look for me food to eat, nothing else I could do, second to that is sleeping at night” (male PWD, FGD)*

### 3.5 Physical environment

The impact of flooding was found to hinder hand hygiene behaviour in several ways. Firstly, floods further limit handwashing by blocking access to water points and submerging water and handwashing facilities. Additionally, most KII participants mentioned floods have led to diversion of humanitarian funds to other pressing needs such as emergency food distribution, also blocking access to routes, preventing NGOs from accessing camps. In 10 FGDs with PWDs, men and women of various age groups, participants consistently reported floods impeding access to handwashing materials such as *Ibrik*, soap and buckets. This is due to a combination of handwashing materials being lost during the floods, as well as lack of access to materials in the camps. The floods have also made it difficult for PWDs to access water points, with mud and flooding restricting movement, as highlighted by women PWD FGD and participants in the elderly women group. Even outside of flooding, PWDs described finding it difficult to access water due to long queues, impeding handwashing behaviour. The floods have also caused latrines to fill up, causing sewage to pour out into the camp.

> *“We never had enough water and unless due to this flooding. If you managed to fetch for two jerrycans cooking and drinking, then you fetched flooded water for other domestic work for washing clothes and bathing.” (young woman, FGD)*

Additionally, during flooding, residents reported commonly using flood water for handwashing. One KII participant attributed this behaviour to shortages of water supplies and the relative proximity and accessibility of flood water. Two FGDs of young and older men also mentioned shortages of water points across all camps, due to the increased number of IDPs due to the continued flooding.

### 3.6 External context

Alongside the direct impacts of flooding, FGD participants across different groups consistently highlighted a lack of active WASH programs in the camps, contributing to previously mentioned barriers with infrastructure and materials, impeding handwashing behaviour. Alongside this, camp residents are experiencing livelihood issues which are further hindering access to materials.

> *“We are unable to buy things, unable to cultivate our farms and our livestock are dead. Hands washing was seriously affected by the flood because water is too much and soaps and bucket for washing are not there.” (older woman FGD)*

Key informants described current camp handwashing and sanitation programs as including garbage collection, handwashing facility installation by both men and women, and latrine and bathroom cleaning. However, informants mostly deemed camp handwashing infrastructure as insufficient due to theft, vandalism and poor maintenance. Specifically, they reported that the cleaners are not equipped with liquid soap and hand gloves and are demotivated due to insufficient pay. Consequently, some latrines and bathrooms are cleaned irregularly, discouraging use. Moreover, FGD and KII participants concurred that the last humanitarian support in terms of *Ibrik*, buckets and soap distribution were in late 2021 or early 2022 (i.e. at least 1.5 years prior to data collection).

### 3.7 Avenues to improve hand hygiene behaviour

Some FGD participants suggested that cleaners should be assigned responsibility for monitoring vandalism and theft, which apparently worked well in another POC, where public handwashing facilities lasted longer. They also suggested the cleaners should be paid well to improve maintenance and that facilities be made more attractive. KII and FGD participants all suggested increasing the working hours for cleaners and hygiene promotors, also suggesting an increase in camp handwashing infrastructure. The FGD with elderly camp residents suggest there should be inclusive hand hygiene messaging in the camps, emphasising the importance of reaching those that are illiterate, children and elderly.

> *“If they construct for us more taps and boreholes it will be good for us. We are also concerned about the materials that organization uses to give us; they are not enough. if each one of us have two buckets at home, I can collect water today and for tomorrow” (female PWD, FGD)*

### 3.8 Summary of behavioural determinants

Below, in table 4, we summarise the key handwashing behavioural determinants among camp residents across the BCD dimensions.

**Table 4.**
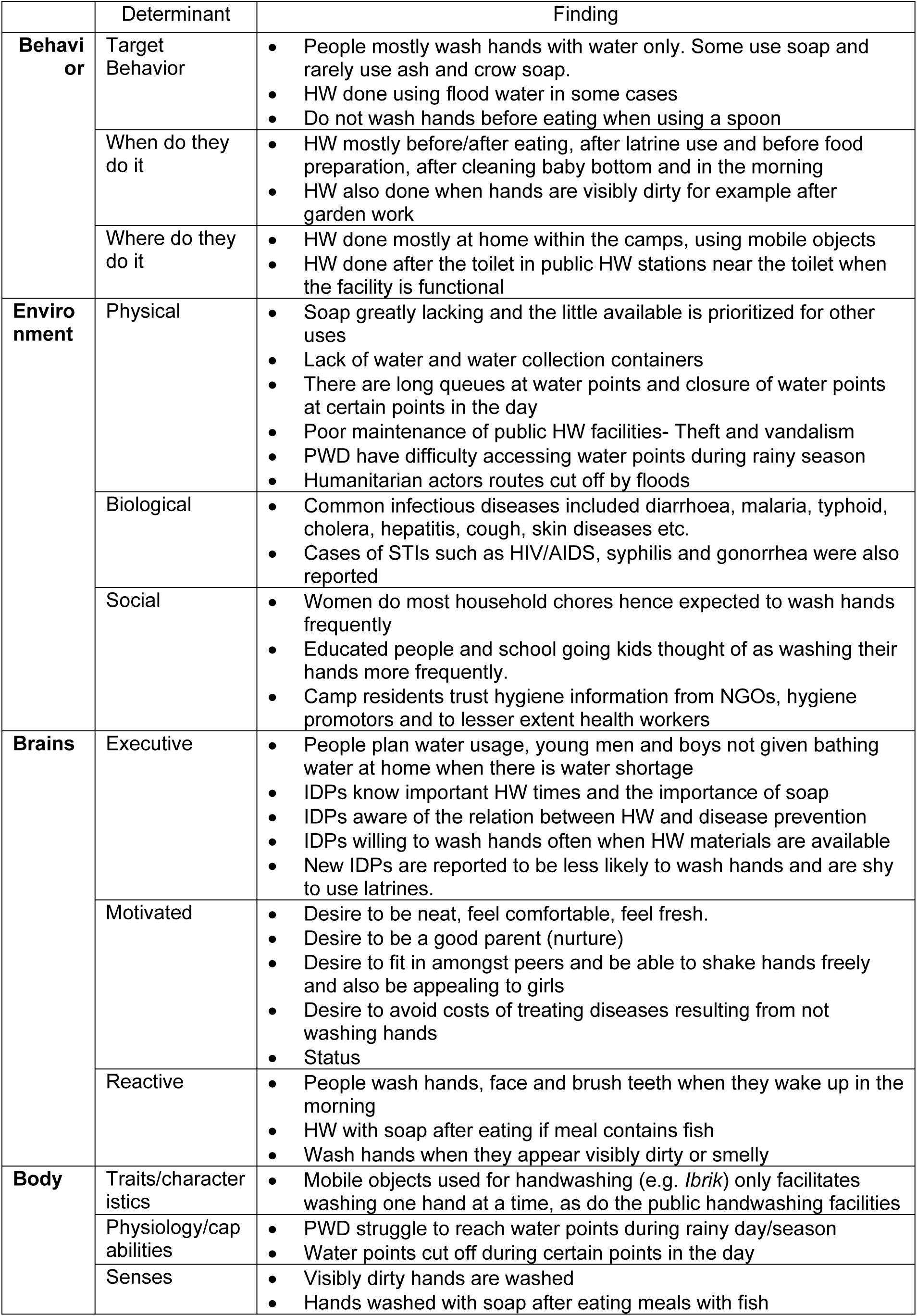
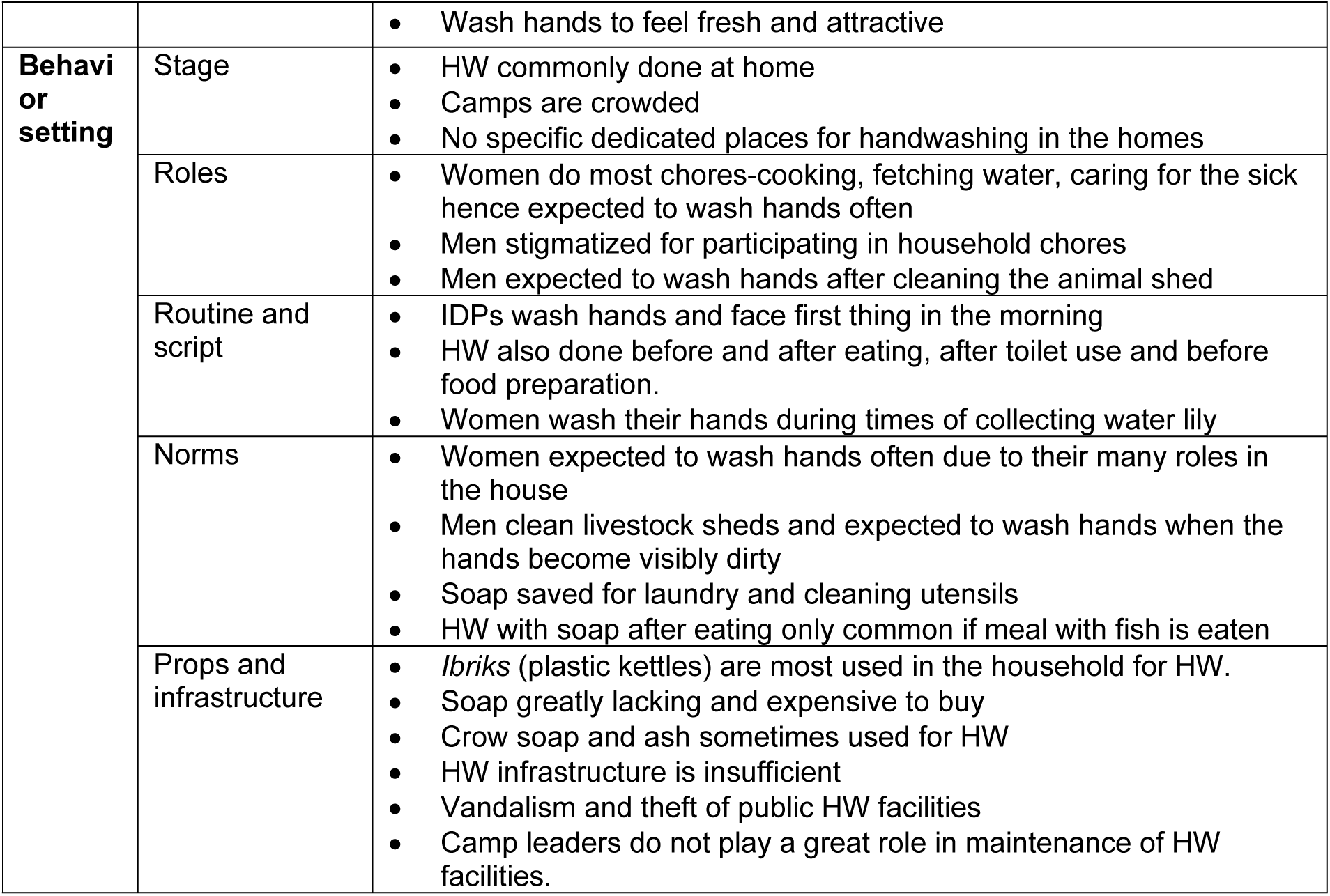
Summary of hand hygiene behavioural determinants among camp residents in four IDP camps in Bentiu, South Sudan.

## 4. Discussion

This study aimed to elucidate the key behavioural determinants of hand hygiene among IDP camp residents in flood-affected areas of South Sudan, using the BCD framework, with a view to co-design a behaviour change intervention. We found that residents generally wash their hands with water only, typically before/after eating and after using the toilet, with women also washing prior to preparing food and after cleaning babies. As soap is generally unavailable, participants prioritise its use for washing clothes and dishes or when their hands are visibly dirty, oily or odorous. Generally, participants understood the role of hand hygiene in preventing diseases and critical times, though a lack of infrastructure and materials is hampering behaviour. Handwashing is motivated by a desire for neatness, comfort, nurture, status, fear of disease, and courtship among young men. Participants reported enjoying the sensory feeling of hand washing with soap.

Women, people who are educated and children are perceived as handwashing more frequently, while PWDs face physical barriers and new residents are perceived as less knowledgeable, hindering hand hygiene. The behaviour setting, characterised by overcrowded, poorly maintained, unattractive, shared public hand hygiene infrastructure with insufficient access to soap and water, as well as the lack of a dedicated handwashing facility in households presents a key barrier. Due to gendered norms, women practice handwashing more frequently and use water for various chores, while norms around handshaking, courtship and expectations for leaders also influences hand hygiene. Overall, we found that the hand hygiene determinants were relatively similar to those identified in non-crisis low-income settings in an integrative systematic review (White et al., 2020a). Finally, we elucidated the multifaceted impacts of flooding, including damage or loss of infrastructure and soap, funding competition halting WASH activities and interactions with unsafe flood water.

### The behaviour setting is a key barrier

Our results indicate that knowledge of importance hand hygiene is not being translated into HW behaviour change, primarily due to insufficient handwashing materials and infrastructure in the camps. This aligns with findings from an integrative systematic review from White et al. (2020a), which found that the knowledge about disease and disease transmission alone, may have limited or no impact on handwashing practices, and that equitable access to materials and infrastructure may be a more important determinant – echoing other studies (Ataiyero et al., 2018; Borg, 2014; Hotopf et al., 2024; WHO, 2021; Zheng et al., 2022). A study by Blum et al. (2019) in IDP camps in the Democratic Republic of Congo also found that the failure to maintain public handwashing stations, the lack of soap distribution and the unavailability of soap due to its cost, was a major barrier of hand hygiene, with handwashing with soap increasing if soap was available. Our study similarly, suggests that improving access to handwashing infrastructure and materials may be most impactful in improving handwashing behaviour. This could be done by providing more public handwashing stations and ensuring there is a system of maintenance established among camp residents, such as through establishing Village Water Committees who oversee WASH related community issues, including maintenance of infrastructure, which proved effective in Tanzania (Kema et al., 2012). Overall, more research on approaches to increasing ownership and maintenance of public facilities is needed, as is advocacy for the prioritization of soap distributions to implementing agencies and donors. This should be in line with existing guidelines, such as the sphere guidelines stating that ‘Appropriate items to support hygiene, health, dignity and well-being are available and used by the affected people.’(Sphere, 2018). Additionally, given the expensive nature of soap and the difficulty of obtaining it, it might be worthwhile to research the feasibility of a soap-making / soapy water -making, which is often less costly, to ensure access to soap in the IDP camps as well as potentially providing income to camp residents, especially women. Whist soap making interventions have been conducted in some settings (Adam et al., 2020; Hetherington et al., 2017; Niyonkuru, 2025), we are unaware of studies evaluating the impact of these activities on handwashing with soap, with studies typically focusing on livelihood impacts - thus further research is warranted. In addition to the public handwashing facilities, interventions should also target household settings to ensure an enabling environment for handwashing, as we found that this is where the majority of handwashing, aside from after the toilet, takes place. Given that access to WASH infrastructure is an equity issue, it is crucial that any intervention embeds an equity focus throughout (Calderón-Villarreal et al., 2022; Hotopf et al., 2024).

### Camp residents have the knowledge and capacity to wash their hands

Our findings are generally consistent with other studies that have found that IDP camp residents have good knowledge of the importance of handwashing in relation to disease prevention, for example in countries such as Iraq (Zangana et al., 2020) and Democratic Republic of Congo (Blum et al., 2019; Claude et al., 2020). However, like in other studies, we found that knowledge alone does not translate into practice, due to other key determinants, such as a lack of infrastructure (White et al., 2020a; WHO, 2021). This emphasises the importance of considering other behavioural determinants when designing and implementing behaviour change interventions. We also found that the new IDP residents were considered to be less knowledgeable and to practice hand hygiene less frequently. Current evidence on the relationship between the duration of camp stay and hand hygiene behaviour is contentious; whilst some literature links the length of stay to a sense complacency, impeding hand hygiene (Biran et al., 2012; Nahimana et al., 2017). Namara et al. (2020) report a significant association between length of stay and household access to a functional handwashing facility, supported by Scobie et al. (2014) who suggest that long-term residents exposed to hygiene promotion demonstrate positive behaviours, including obtaining handwashing facilities. Therefore, we recommend that future interventions target new IDPs arriving at camps for behaviour change messaging and activities.

### Gender plays a key role in hand washing behaviour

There is a gendered difference in hand hygiene, largely due to strong social norms that dictate gender roles within the camps in South Sudan and the country more broadly. Household water collection, childcare, nursing the sick, collecting water lilies, food preparation and cleaning dishes fall under household chores acceptable to only women, with men stigmatised for doing these activities. As result of these gender inequities, women reported practicing hand hygiene more regularly than the men. These results resonate with findings from the review by White et al. (2020a) which found that being female had a mixed but positive association with handwashing. Another study conducted in a university in Ghana found handwashing behaviour was strongly gendered, with women more likely to wash their hands in general, and wash both hands, compared to men (Mariwah et al., 2012). However, a study in Sierra Leone reported that whilst the traditional roles of women (e.g., childcare and food preparation) motivate hand hygiene intention and present key handwashing points for women, gendered power inequities can impede access to soap, and thus hand hygiene, as women may encounter fewer opportunities to purchase soap (Lanfer & Reifegerste, 2021). Our study also highlighted that young men wash their hands more whilst courting women, as cleanliness is deemed attractive and it was reported that school going children were perceived to be more receptive to handwashing, regardless of gender. Therefore, behaviour change messaging and activities should consider gender, perhaps centring around status (men) and nurture (women) and comfort and social affiliation, with a focus on engaging men and having women and school children championing the cause. Finally, our study highlighted potential inequities in the gender of WASH professionals working in the camps, indicated by the research team only being able to recruit one female key informant. Implementers should strive for gender balance in their teams generally, particularly among those involved in the design and implementation of WASH interventions (WaterAid, 2023).

### Sensory perception is a key behavioural determinant

Participants consistently highlighted the role of sense in driving hand hygiene behaviour; feelings of freshness and cleanliness drove behaviour, as did undesirable sensations such hands being visibly oily, dirty or unpleasant olfaction associated with certain foods, wherein soap was prioritised. These findings are supported by existing evidence which highlights the role of senses, namely feeling or being perceived as dirty, in driving hand hygiene (Curtis et al., 2009; White et al., 2022; Zangana et al., 2020). In an 11-country review, Curtis et al. (2009) elucidated important motivations for handwashing, including disgust (e.g., due to contamination with fish or bodily fluids, which influences social status and affiliation), comfort (feeling fresh and clean, which is associated with confidence, including during social contacts), nurture (in terms of being a caring mother and ensuring the health of one’s child) and attraction (cleanliness associated with the ability to attract “high-value mates”), all also reflected in our findings. The strong aversion to visible dirt and desire for feeling clean may be contributing to the relatively low use of ash among residents, as it does not leave hands feeling or looking clean (White, 2020b).

Despite participants demonstrating strong knowledge of the role of hand hygiene in disease prevention, the fact that the use of soap was mentioned more often in circumstances where hands were dirtied after eating, above prior to feeding one’s child and after cleaning a child’s bottom, also suggests a knowledge gap about critical points (GHP, 2020). Future interventions might focus on promoting the prioritisation of soap at these critical times, and using appropriate technique, through drawing on the motives of comfort and disgust, and social norms. This could also involve promoting hand washing with low-cost soap solutions, such as soapy water, after eating food to achieve a sense of cleanliness whilst saving soap for critical times (Amin et al., 2014; MacLeod et al., 2023; WHO, 2022b).

### Multifaceted impact of flooding

Flooding is reportedly impeding IDP residents’ hand hygiene behaviour and possibly driving outbreaks of respiratory and diarrhoeal diseases through numerous intersecting pathways (**Error! Reference source not found.**). Firstly, access to WASH infrastructure, including water points and handwashing facilities, is reportedly being compromised by submersion and destruction associated with floods, with PWDs disproportionately impacted. Moreover, residents reported losing mobile objects (e.g., *ibriks*) and handwashing materials during flooding events. Alongside this, respondents assert that the diversion of funds to other more acute programming, and the influx of more IDPs, is further reducing WASH programming capacities and straining limited resources. Together, persons affected feel that these factors are impeding hand hygiene behaviour. These findings are consistent with the literature; a study in camps in Northeast Nigeria found flooding was associated with destruction of WASH infrastructure and the overburdening of WASH services due to incoming IDPs (Jaber et al., 2023), with the latter also captured in Kenya (Mahamud et al., 2011). It is also widely acknowledged that people with disabilities face health inequities, partially due to physical and social barriers accessing essential services, including WASH infrastructure and programming, and are disproportionately impacted by extreme weather events, including flooding (Stein et al., 2024; WHO, 2022c). Additionally, flooding can introduce pathogens through latrines overflowing and contaminating surface water, which may lead to residents using contaminated flooding water for handwashing and playing. Indeed, a systematic review on the epidemiology of floods in sub-Saharan Africa reported that 90% of studies found an increased probability of being affected by diseases, including cholera, whilst a study in South Africa found that interaction with contaminated surface water was a key WASH issue following floods (Suhr & Steinert, 2022; Tshuma et al., 2023). Post-flood outbreaks of diarrhoeal and respiratory diseases have also been recorded in studies in South Sudan, specifically (Desai An Fau - Mohareb et al., 2022). Such outbreaks are also associated with higher morbidity and mortality rates in IDP settings, due to numerous intersecting inequalities, such as overcrowding, inadequate access to essential services and food insecurity (Cantor et al., 2021; Gichunge et al., 2020; Thorseth et al., 2021).

Therefore, we suggest that implementers consider flood risk when building WASH infrastructure and ensure that PWDs and other marginalised groups are actively engaged when planning and implementing WASH programming to promote equitable access. Additionally, we need to invest in researching approaches to integrating behaviour change messaging with other crisis-response activities and incorporating system strengthening activities, to support the establishment of sustainable, resilient systems which can respond to future crises (Bogale et al., 2024; Esteves Mills et al., 2023). This might include strengthening multisectoral collaboration in camps, supporting local markets to respond to demand (e.g., training artisans to build and repair handwashing facilities), strengthening community health workers’ capacity, contributing to national monitoring systems and using evidence to advocate for increased WASH prioritisation (Bogale et al., 2024; Durrance-Bagale et al., 2020; Hotopf et al., 2023). Finally, we need to invest in in research and messaging on the safety of using flood water for hygiene practices.

### Strengths and limitations

This study contributes to the limited pool of evidence on hand hygiene behavioural determinants within flood affected settings among IDPs, who have historically been under prioritised in research (Swartz et al., 2023). We used participatory research to centre the lived experience of persons affected, and actively engaged marginalised populations, including PWDs and women. Finally, through triangulating multiple methodological approaches and participant types, we enhanced the credibility, dependability and confirmability of our findings.

Despite these strengths, there are numerous limitations which must be considered. Firstly, the handwashing demonstration and surveys are both susceptible to observation bias and recall bias respectively, as well as social desirability bias. Moreover, whilst we intended to conduct two FGDs with men and women with disabilities, it proved difficult to recruit participants and ended up only conducting one with each gender. However, we generally found that findings echoed those of groups with participants without disabilities, suggesting that saturation may still have been reached, with no major implications on the conclusions drawn. Similarly, whilst we had planned for a gender balance in key informants, we were only able to engage one female participant, potentially introducing bias.

There were also numerous challenges faced during data collection, which were associated with working in hard-to-reach settings and changes in budgets due to funding cuts. This includes poor communication due to network issues and using data collectors with whom the research team did not have a pre-established relationship with, resulting in issues during data collection. Firstly, the camp tours were not conducted as intended; we had planned to systematically record information on all WASH infrastructure observed in the tours (sanitation, water, and hygiene), however sanitation information was not collected and the rest of the data were lost when the tablets were lost in transit, which limits our ability to quantify access and coverage. While we don’t have reliable data on coverage of facilities, we know that there were some facilities in all the camps but that these were not considered sufficient by FGD participants and KIIs. Additionally, data from one of the camp leader interviews went missing. These interviews were brief, and in retrospect, we could have explored more barriers to hand hygiene from the camp managers’ perspectives. There were also issues with data collection for some of the participatory activities in the FGDs; for instance, sufficient data was only collected for two of the motives exercises and data from the water prioritisation exercise was also incomplete, due to the loss of notes by data collectors. In hindsight, the SOPs and training should have been more specific and emphasized that audio recordings alone were not sufficient for capturing the results of the participatory exercises. Finally, it is important to note that the study was conducted in four IDP camps in South Sudan and whilst our study contributes to the small pool of literature on hand hygiene in IDP camps impacted by flooding, the results are not necessarily generalisable to other settings. Linked to this, is the limitation that our study was conducted in camps where soap, which is critical for handwashing, was largely unavailable, which limits the determinants that could be identified.

## Conclusion

Our research demonstrates that despite knowledge of hand hygiene’s role in disease prevention, key techniques and some of the critical times, residents of Bentiu IDP camps are not frequently practicing handwashing with soap. Consequently, residents are at heightened risk of contracting and transmitting infectious diseases. Handwashing practice is being hindered by insufficient access to infrastructure and materials, with flooding amplifying challenges. Whilst we identify several other determinants which could be targeted to implement person-centred evidence-based behaviour change interventions, their efficacy will be limited unless the behaviour setting is addressed and persons affected have equitable access to WASH infrastructure and materials.

## Data Availability

The datasets analysed during the current study are available from the corresponding author upon request and any further clarifications are available on request.

## Acknowledgements

We would like to thank all participants of this research, without whom this study would not have been possible. We are also thankful to the research assistants: Deng Gatliah, Marko Gatjang, Nyatinga Michael, Tabitha Nyachiang. We are also grateful to Sian White who provided suggestions during the protocol development phase, Robert Aunger at LSHTM for his valuable inputs on an early version of this manuscript and Tom Handzel and Christina Craig from the CDC for their helpful comments and suggestions on a later version.

## Notes

### Competing Interest Statement

The authors have declared no competing interest.

### Funding Statement

This publication is supported by the Centers for Disease Control and Prevention of the U.S. Department of Health and Human Services (HHS) as part of financial assistance award 5U01GH002319. The contents are those of the author(s) and do not necessarily represent the official views of, nor an endorsement, by CDC/HHS, or the U.S. Government.

### Author Declarations

Ethical approval was obtained from the LSHTM ethics committee (Ref: 28372) and the South Sudan Ministry of Health (MOH/RERB/A/05/2023).

